# Polycystic Ovary Syndrome in Medical Students: A Cross-Sectional Study of the Academic and Psychosocial Impact

**DOI:** 10.1101/2025.07.03.25330829

**Authors:** Ashini A. Patel, Caitlyn R. Curley, Ganesh Chilukuri, Hannah Lee, Samantha Snyder, Vivek Joshi

**Author notes:** Ashini Patel –. Caitlyn Curley –. Ganesh Chilukuri –. Hannah Lee –.

## Abstract

**Background:** Polycystic Ovary Syndrome (PCOS) is a prevalent endocrine disorder affecting up to 13% of reproductive-aged females, often accompanied by physical, metabolic, and psychological challenges. Despite the demanding nature of medical education, limited research explores the burden of PCOS among medical students.

**Objective:** To evaluate the academic, emotional, and psychosocial impact of PCOS on medical students across four U.S. medical schools.

**Methods:** A cross-sectional survey was distributed to assigned female at birth medical students aged 18 and older. The questionnaire assessed PCOS diagnosis and treatment, academic performance, symptom burden, emotional well-being, and coping mechanisms. Descriptive statistics and Pearson correlation analyses were performed.

**Results:** Of the 380 respondents (mean age 25.9), 30.3% reported a PCOS diagnosis—more than double the estimated global prevalence. PCOS was associated with academic stress (34.0%) and reduced work productivity due to emotional distress (70.2%). Symptom burden included menstrual irregularities, weight concerns, bloating, and acne. Emotional impacts were significant: 49.3% reported feeling unattractive, 50.7% worried about infertility, and 50.7% experienced self-blame. While many students demonstrated resilience and adopted positive coping strategies, a notable proportion reported emotional exhaustion and giving up efforts to manage the condition.

**Conclusion:** Medical students with PCOS experience significant emotional and academic challenges, compounded by the rigors of their training. The prevalence and impact of PCOS in this population highlight the need for targeted institutional support, improved awareness, and mental health resources to promote student well-being and success.

## Introduction

Polycystic Ovary Syndrome (PCOS) is a prevalent endocrine disorder affecting up to 13% of reproductive age females worldwide [1]. Characterized by hyperandrogenism, ovulatory dysfunction, and polycystic ovarian morphology, PCOS presents a complex interplay of metabolic, reproductive, and psychological challenges [1,2]. Despite its significant impact on physical and mental health, research on PCOS remains underfunded and limited, particularly regarding its effects on specific subpopulations. While studies have examined the psychological and social implications of PCOS in the general population, there is a paucity of research investigating its influence on medical students—a group uniquely positioned at the intersection of academic rigor and healthcare awareness.

Medical students experience heightened stress levels due to demanding coursework, clinical responsibilities, and performance expectations [3]. When coupled with the symptoms of PCOS, such as menstrual irregularities, weight fluctuations, fatigue, and mental health disturbances, their academic performance and overall well-being may be further compromised. Understanding how PCOS affects future physician generations is crucial for identifying tailored interventions that support their educational and professional development.

Emerging evidence suggests a higher prevalence of PCOS among medical students than the general population. One study reported that 15% of 231 medical students surveyed exhibited symptoms consistent with PCOS, indicating a potentially disproportionate burden within this cohort [4]. Moreover, qualitative interviews highlight increased stress, anxiety, and challenges in balancing academic responsibilities among students diagnosed with PCOS. Another study demonstrated that adolescents with PCOS features exhibit metabolic risks, emphasizing the disorder’s long-term health implications [5]. However, there is a critical gap in literature exploring the specific experiences of medical students with PCOS and the ways in which the condition influences their academic pursuits.

Our study seeks to address this gap by investigating the experiences of medical students diagnosed with PCOS across four U.S. medical schools. Through a self-administered survey targeting biologically assigned females (AFAB) aged 18 and older, we aim to assess the impact of PCOS on academic performance, mental health, and overall quality of life. Given the established association between PCOS and mental health disorders, our research endeavors to provide data-driven insights that can inform institutional policies and the development of tailored support systems for affected students. By enhancing awareness and fostering institutional accommodations, we hope to improve the academic success and well-being of medical students with PCOS. Research indicates that early intervention in managing PCOS can significantly enhance fertility outcomes. A systematic review and meta-analysis concluded that first-line pharmacological treatments, especially when combining insulin sensitizers with conventional ovulation stimulants, effectively improve clinical pregnancy rates in women with PCOS-related infertility [6]. This underscores the importance of prompt diagnosis and tailored treatment plans for women with PCOS, particularly those navigating the rigorous demands of medical careers.

## Methods

### Study Design and Participants

This cross-sectional study was conducted between July 2024 and January 2025 across four accredited U.S. medical schools. Participants were recruited via institutional email listservs, student organization announcements, and social media groups. Participation was voluntary, and informed consent was obtained electronically before the start of the survey.

### Survey Instrument

Data was collected using a self-administered, anonymous online questionnaire specifically developed for this study. The questionnaire was designed following a review of existing instruments assessing PCOS-related quality of life, coping mechanisms, and academic performance, and was pilot-evaluated on a small sample of medical students (n=10) to ensure clarity and relevance before broader dissemination. The survey included five major sections: demographics (age, ethnicity, academic year, and curriculum type, such as self-directed learning [SDL], lecture-based, or problem-based learning [PBL]); PCOS diagnosis and treatment (questions regarding clinical diagnosis and treatment modalities, including medical, surgical, or alternative therapies); academic performance and stress (assessing perceived academic satisfaction and stress attributed to PCOS); quality of life (measuring the frequency of symptoms such as headaches, bloating, menstrual cramps, weight concerns, excess hair growth, and acne, as well as general health, emotional well-being, physical function, and work productivity); and coping strategies (capturing the extent of engagement in supportive activities, lifestyle adjustments, denial, self-blame, or religious practices). Responses were collected using Likert-type scales appropriate to each section, typically ranging from 1 (‘Always’) to 5 (‘Never’) or using similar ordinal formats.

### Data Collection

The survey was hosted on a secure online platform (Qualtrics®), and responses were collected anonymously. IP addresses were not stored, ensuring participant confidentiality. Completion of the survey took approximately 10–15 minutes. Data collection continued until a sample size sufficient for meaningful statistical analysis was achieved.

### Statistical Analysis

Descriptive statistics, including means, standard deviations, frequencies, and percentages, were used to summarize demographic characteristics, prevalence of PCOS diagnosis, treatment patterns, academic performance, quality of life indicators, and coping strategies. Bivariate analyses, including Pearson correlation coefficients, were conducted to explore associations between PCOS status, symptom burden, academic stress, coping mechanisms, and emotional well-being. A significance threshold of p < 0.05 was used for all inferential statistics. All analyses were performed using IBM SPSS Statistics version 25.0.

### Ethical Considerations

The study was approved by the Institutional Review Boards (IRBs) of all participating institutions. Participation was voluntary, and no identifying information was collected. Participants were informed about the study’s objectives, the voluntary nature of participation, and their right to withdraw at any time without any penalty.

## Results

**Table 1.** Demographic and Clinical Characteristics of Participants (N = 380)

| Variable | Mean ± SD /<br>Range | Frequency (%) |
| --- | --- | --- |
| Age (years) | 25.90 ± 2.66 | Most common: 25 (18.2%), 27 (17.1%), 28 (18.4%) |
| Diagnosed with PCOS |  | 30.3% |
| PCOS Treatment Type |  | Medical: 86.5%<br>Alternative: 13.5% |

**Table 2.** Academic Performance, Quality of Life, and Emotional Health.

| Category | Symptom | Mean ± SD | Most Common Response (%) |
| --- | --- | --- | --- |
| Academic Impact | Academic Satisfaction | 1.96 ± 0.58 | Satisfactory: 66.2% |
| | Curriculum Type | 2.26 ± 1.16 | SDL: 38.8% |
| | Stress due to PCOS | 2.06 ± 0.86 | No: 39.6%, Yes: 34.0% |
| Quality of Life | Headaches | 4.00 ± 0.85 | Sometimes: 43.3%,<br>Never: 31.1% |
| | Abdominal Bloating | 3.49 ± 1.12 | Most of the time: 29.8% |
| | Menstrual Cramps | 3.13 ± 1.62 | Always: 28.0% |
| | Irregular Cycles | 2.79 ± 1.55 | Always: 26.9%, Most: 29.3% |
| Emotional Health | Concern about Weight | 2.15 ± 1.50 | Always: 57.8% |
| | Frustration Losing Weight | 2.13 ± 1.48 | Always: 53.3% |
| | Excess Hair Concerns (Q15) | 2.73 ± 1.35 | Most: 35.1% |
| | Acne Concerns | 2.40 ± 1.59 | Always: 45.1% |
| | Feeling Unattractive | 2.30 ± 1.46 | Always: 49.3% |
| | Infertility Concerns | 1.49 ± 0.50 | Always: 50.7% |
| | Cancer Concerns | 4.01 ± 1.13 | Never: 51.2% |
| General Health | General Health Rating | 3.21 ± 0.97 | Half the time: 44.9% |
|  | Anxiety About PCOS | 3.12 ± 1.24 | Half the time: 42.0% |

**Table 3.** Physical Function, Work Productivity, and Coping Strategies.

| Category | Item / Strategy | Mean ± SD | Most Common Response (%) |
| --- | --- | --- | --- |
| Physical Function | Limited in Moderate Activity (Q24) | 2.67 ± 0.47 | Not at all: 67.3% |
|  | Climb Several Flights of Stairs (Q25) | 2.61 ± 0.49 | Not at all: 61.5% |
| Productivity | Less Work Due to Physical Demands (Q26) | 1.64 ± 0.48 | Yes: 35.9% |
|  | Less Work Due to Emotional Stress (Q27) | 1.30 ± 0.46 | Yes: 70.2% |
| Coping | Learning to Live with PCOS (Q38) | 3.10 ± 1.00 | A lot: 50.7% |
|  | Turning to Activities (Q32) | 2.30 ± 1.08 | None: 30.9% |
|  | Self-Blame (Q39) | 1.92 ± 0.94 | A lot: 50.7% |
|  | Use of Substances (Q34) | 1.41 ± 0.70 | Never: 71.2% |
|  | Seeking Help/Advice (Q35) | 2.40 ± 1.05 | Medium time: 49.3% |
|  | Gave up Dealing with PCOS (Q36) | 1.57 ± 0.74 | Yes: 31.7% |

### Participant Characteristics

A total of 380 participants were included in the study, with a mean age of 25.90 years (SD = 2.66). Among them, 30.3% reported having been diagnosed with PCOS, while 69.7% had not. Of those diagnosed, 86.5% were receiving medical treatment, and 13.5% were using alternative therapies.

### Academic Performance and Stress

Regarding academic performance, 19.0% of participants reported high satisfaction, 66.2% were satisfied, and 14.8% reported less satisfaction. Participants followed different curriculum types, including self-directed learning (38.8%), lecture-based (15.8%), and problem-based learning (26.1%). Notably, 34.0% of participants reported experiencing stress about their academic performance being affected by PCOS, while 39.6% did not.

### Quality of Life and Symptom Burden

PCOS symptoms affected participants’ quality of life in multiple ways. The most frequently reported symptoms included headaches (43.3% sometimes), abdominal bloating (29.8% most of the time, 39.3% sometimes), and menstrual cramps (28.0% always, 14.8% half the time). Additionally, 26.9% reported experiencing irregular menstrual cycles always, while 29.3% reported this occurring most of the time. Weight concerns were prevalent, with 57.8% always feeling concerned about being overweight and 53.3% reporting frustration in attempting to lose weight. Excess facial and body hair was also a concern, with 35.1% reporting the need for hair removal most of the time.

### Emotional and Psychological Impact

Participants expressed significant emotional distress related to PCOS. Approximately 49.3% always felt unattractive due to their weight, body hair, or acne, while 29.0% reported feeling fearful of how others perceived their appearance. Concerns regarding future infertility were reported by 50.7% of participants, while 42.0% felt anxious about their PCOS half the time. Additionally, 51.2% never felt frightened about developing cancer due to PCOS.

### Physical Function and Work Productivity

Regarding physical activity, 67.3% of participants reported no limitations in moderate activities, while 61.5% were able to climb several flights of stairs without difficulty. Work productivity was affected by physical and emotional demands, with 35.9% reporting accomplishing less work due to physical demands and 70.2% due to emotional stress.

### Coping Strategies

Participants adopted various coping strategies to manage PCOS. While 50.7% reported learning to live with PCOS a lot of the time, 66.8% had not turned to alcohol or substances to cope. Instead, 28.2% engaged in activities such as exercise, cooking, or social media as a distraction. Seeking support from others was another strategy, with 49.3% doing so for at least a medium amount of time. However, 31.7% reported having given up trying to deal with PCOS, and 50.7% engaged in self-blame frequently.

## Discussion

This study offers insights into the experiences of medical students diagnosed with PCOS, shedding light on how it impacts their academic performance, mental health, and overall quality of life. The demographics provide context for how we interpret this data. With a mean age of 25.9 years old, this cohort aligns with the typical age group affected by PCOS. More notably, 30.3% of the participants reported having PCOS, significantly exceeding the global prevalence of approximately 11-13% [1-3]. This suggests a disproportionate burden among this population and emphasizes the need for targeted institutional support.

The symptom burden reported by students diagnosed with PCOS indicates a multifaceted impact on their daily well-being. Common complaints such as abdominal bloating, menstrual cramps, and irregular cycles were reported with high frequency, highlighting the chronic nature of PCOS symptomatology. Furthermore, the was significant concern about weight with 57.8% reporting that they were always concerned and 53.3% reporting frustration with weight loss efforts. This aligns with well-documented challenges related to insulin resistance and metabolic dysfunction in PCOS patients [12-13]. The reports of physical discomfort and body image concerns related to hirsutism, acne, and bloating further negatively impact quality of life and contribute to emotional distress. Similar findings showed that women with PCOS have impaired health-related quality of life, particularly driven by symptoms related to physical appearance and menstrual irregularities, which frequently contribute to psychological distress [7]. The stress burden can impact their academic performance. In our study, 34% of respondents reported experiencing stress specifically related to concerns about how PCOS was impacting their academic performance, which has been corroborated with previous studies highlighting the increased academic and emotional strain experienced by medical students with PCOS [12-14].

Emotional distress was also highly prevalent among participants diagnosed with PCOS. Nearly half (49.3%) reported persistent feelings of unattractiveness related to symptoms like weight issues, acne, or hirsutism, which are manifestations often associated with hyperandrogenism [3,5]. Additionally, 50.7% expressed anxiety about future infertility, highlighting the specific reproductive concerns within this group. PCOS is one of the leading causes of female infertility, as irregular ovulation or anovulation can significantly reduce the chances of conception [15].

This can be particularly distressing for female physicians, who often face delays in family planning due to the demands of medical training and professional responsibilities. Given that many female physicians postpone family planning due to career demands [9], the interplay between professional goals and reproductive health challenges in women with PCOS deserves particular attention. Prior research supports that infertility-related stress is notably elevated in women with PCOS, further amplifying anxiety in an already high-pressure environment like medical training [10,14].

Regarding physical function and work productivity, more than two-thirds indicated no limitations with moderate activity related to their PCOS. However, 70.2% of participants reported accomplishing less work due to emotional demands. Huddleston et al. (2024) reported similar findings, with 51.5% of participants stating that PCOS interfered with their work performance, primarily due to symptoms of anxiety and depression. The disparity between physical capability and reduced productivity highlights how emotional health plays a critical role in academic and professional performance [14].

Finally, students reported a variety of ways they coped in response to their PCOS symptoms. Of note, just over half (50.7%) reported learning to manage and accept the condition over time. Most students did not turn to maladaptive behaviors like substance use, and many cited positive outlets such as hobbies or social activities like spending time with friends. These strategies reflect a sense of resilience and the ability to adapt despite ongoing challenges. However, these findings also showed that 31.7% of students said they had given up trying to manage their PCOS and 50.7% admitted blaming themselves for their symptoms [8-10]. These responses point to internalization of the stigma around PCOS and also to feelings of burnout that come from managing chronic conditions, while also managing the rigorous demands of medical training. Overall, this highlights a potential gap in accessible mental health resources tailored to students dealing with long-term health issues.

Given the high prevalence and multifaceted burden of PCOS among medical students, medical institutions must consider implementing targeted support mechanisms. These could include improved access to on-campus mental health services with providers knowledgeable about chronic health conditions like PCOS. Academic accommodations such as flexible scheduling, extensions on coursework, or medical leave policies should be clearly outlined and accessible to affected students. Institutions could also consider offering peer-support groups or wellness workshops specifically addressing reproductive and hormonal health13. Finally, training faculty and academic advisors to recognize and respond to the unique challenges faced by students with PCOS may foster a more inclusive educational environment [15]. Integrating such resources within medical education infrastructure can help mitigate the emotional and academic toll of PCOS, ultimately promoting student retention, well-being, and professional success. Further studies with more diverse samples and longitudinal follow-up would be beneficial to build on the findings.

## Conclusion

This study underscores the complex burden PCOS places on medical students, highlighting a prevalence notably higher than that seen in the general population. Beyond the physical symptoms, many students reported significant emotional strain, including anxiety, concerns about fertility, and struggles with body image. Although most participants continued to function well physically and expressed general academic satisfaction, the emotional weight of managing a chronic condition alongside the pressures of medical training was evident. Some students demonstrated strong coping skills and resilience, but others described emotional exhaustion, self-blame, and a sense of discouragement. These findings point to a clear need for medical schools to offer more targeted and accessible support systems for students managing PCOS.

## Limitations

The main limitation to this study is, the study relies on self-reported data, which introduces the potential for recall bias and social desirability bias, especially regarding sensitive topics such as body image and infertility concerns. The other limitation is, the cross-sectional design precludes any temporal or causal interpretations between PCOS and academic or emotional outcomes. Lastly, there is a risk of selection bias, as students with more severe symptoms may have been more motivated to respond, possibly inflating prevalence and impact estimates.

## Contribution

The authors AP, VJ, GC, CC, HL and SS conceptualized the study. VJ, CC, GC, SS, HL and AP wrote the first draft. AP, VJ, GC, CC, HL and SS analyzed and interpreted the data. All the authors contributed to its administration, discussion, conclusion, and critical revision. All authors approve the final manuscript for submission.

## Data availability

The data generated during the research and analysis are not available publicly but are available from the corresponding author on a reasonable request.

